# A graphical user interface for editing keypoints from human pose estimation algorithms

**DOI:** 10.1101/2025.11.21.25340755

**Authors:** Zechen Yang, Jan Stenum, Rini Varghese, Ryan T. Roemmich

## Abstract

Human pose estimation algorithms offer exciting potential for performing markerless kinematic tracking of human movement using only simple videos recorded from common household devices (e.g., smartphones, tablets, webcams). However, these algorithms can be limited by mis-tracking or failure to track certain “keypoints” (i.e., digital markers) associated with specific anatomical locations. These tracking errors can be caused by a wide variety of factors, including hallucinations, occlusions, and noise (among others). There is a need for new tools capable of editing the keypoints resulting from applications of human pose estimation algorithms to ensure accurate kinematic tracking throughout the video of interest. Here, we developed and tested a new pose editor tool that provides a graphical user interface (GUI) enabling users to 1) correct mis-tracked or missing keypoints by manually relocating them to their optimal visual locations directly on the video and 2) run pose estimation directly within the GUI for improved accessibility. We tested the tool on a previous dataset of pose estimation-based keypoints of persons with stroke walking on a treadmill. We observed significant improvements in the mean absolute error between keypoint-based kinematics and ground-truth motion capture data and significant improvements in the correlations between the time series data of the keypoint-based kinematics and the ground-truth motion capture data. Finally, we also observed that the time spent performing manual editing showed a significant positive relationship with the degree of improvement in the kinematic estimates. We have made all software freely available at our GitHub repository: https://github.com/JeffZC/pose-editor.

## INTRODUCTION

Pose estimation algorithms have emerged as powerful technologies for measuring human movement kinematics from simple digital videos [1–9]. These algorithms leverage computer vision to identify and track anatomical landmarks on people within a video via automated application of virtual markers (i.e., “keypoints”), providing a markerless motion capture approach that can be implemented using common household video recording devices (e.g., smartphones, tablets, webcams). Due to their comparatively minimal requirements of time, cost, technical expertise, and hardware, these technologies have the potential to expand access to motion capture far beyond the traditional laboratory setting [10]. Indeed, pose estimation algorithms have been leveraged to measure and assess a wide variety of different human behaviors, including walking [11–17], hand and finger movements [18–21], and other movements of the upper limbs [19,22].

Despite their promise for improving accessibility to motion capture, video-based pose estimation algorithms are susceptible to several different issues that can negatively impact tracking quality [10,23,24]. As examples, pose estimation algorithms are prone to hallucinations (where inanimate objects are tracked as humans or identified body orientations are physically impossible), occlusions (where objects within the video occlude the view of certain anatomical locations, causing difficulty with landmark identification and tracking), and noise (where tracking of specific anatomical locations may be inaccurate due to poor image quality, poor lighting, or the clothing of the person in the video). Additionally, many users rely heavily on pretrained pose estimation algorithms that are often trained on data from limited contexts than in which they might be applied [2,3,5–9]. As one example, we recently showed that OpenPose (a pretrained algorithm for human pose estimation [2,3]) would occasionally show aberrant or missing tracking of the ankle and foot when participants with stroke walked on a treadmill (Figures 1A and 1B) [13]. While many pretrained algorithms provide an output video that shows an overlay of tracked keypoints [2,4,9], the lack of capabilities to edit or reposition the keypoints when errors occur leaves users with little control over data quality, limiting the broader adoption of these tools to varied contexts.

**Figure 1.**
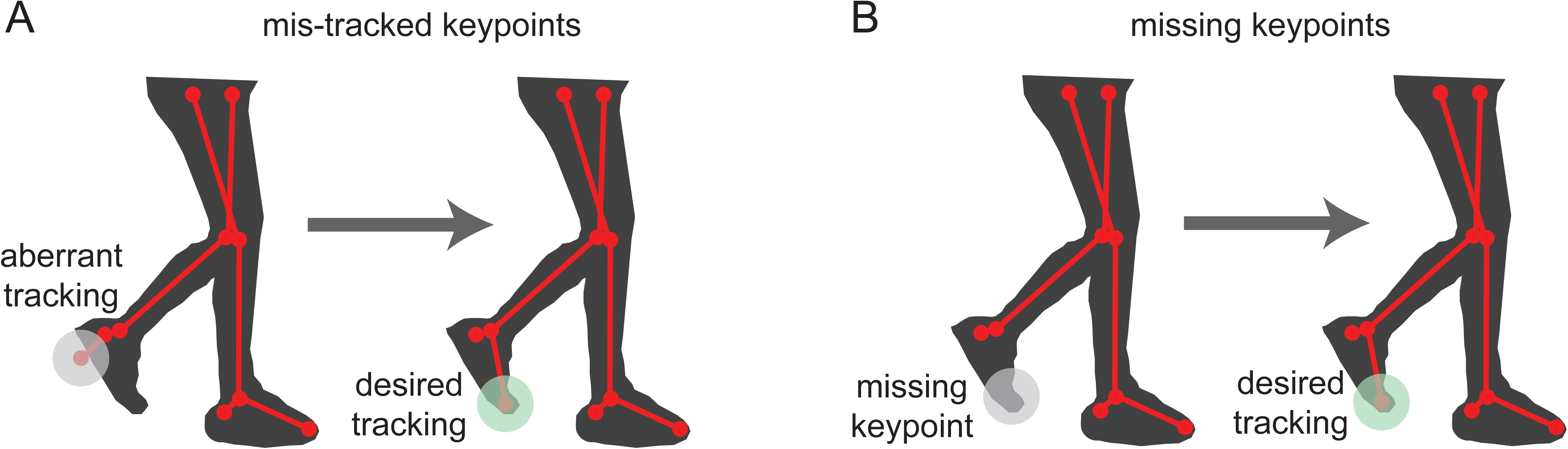
Examples of A) mis-tracked and B) missing keypoints that result in a need for manual editing of pose estimation-based keypoints.

Here, we addressed the need for an accessible, user-friendly tool that can allow users to improve the accuracy of their pose estimation analyses by manually correcting inaccurate or missing keypoints via post hoc intervention. We developed a graphical user interface (GUI) “pose editor” where the user could click and drag the keypoints to correct the tracking frame-by-frame, making the keypoints as close to their true anatomical locations as possible. We also considered it helpful to include a mechanism for applying a pretrained pose estimation algorithm (Google Mediapipe [9,25]) directly within the GUI so that a user can both apply pose estimation and then inspect and/or correct the keypoints within the same application. Finally, we tested the pose editor on the treadmill walking data from persons post-stroke described above to demonstrate a use case where the editor may improve the accuracy of the pose estimation keypoints.

We hypothesized that editing the keypoints with the pose editor would result in stronger agreement and lower error when compared to the ground-truth, three-dimensional motion capture in ankle kinematics during gait in persons’ post-stroke. We also expected that poor quality videos would require more editing time, and that greater editing time will be associated with larger improvements in tracking accuracy after accounting for baseline video quality. All necessary code and instructions for the pose editor GUI are freely available at https://github.com/JeffZC/pose-editor.

## METHODS

### Pose editor GUI

#### Loading existing pose estimation keypoint data

Our pose editor lets the user manually modify frame-by-frame keypoint data from the commonly used human pose estimation algorithms OpenPose (BODY_25 model; [2,3]) and Mediapipe/BlazePose (Mediapipe holistic model; [9]). The pose editor uses a simplified model containing the 21 shared keypoints between the OpenPose BODY_25 and Mediapipe holistic models: nose, left and right eye, ear, shoulder, elbow, wrist, hip, knee, ankle, heel, foot (big toe). This model omits the jugular notch, the center of the pelvis, and the fifth left and right metatarsal of the OpenPose BODY_25 model; and the eyes, mouth, and fingers of the MediaPipe holistic model. A simplified depiction of the GUI before and after manual editing of the pose estimation keypoints is shown in Figure 2 (actual screenshot of the GUI before and after editing is shown in Supplementary Figure 1).

**Figure 2.**
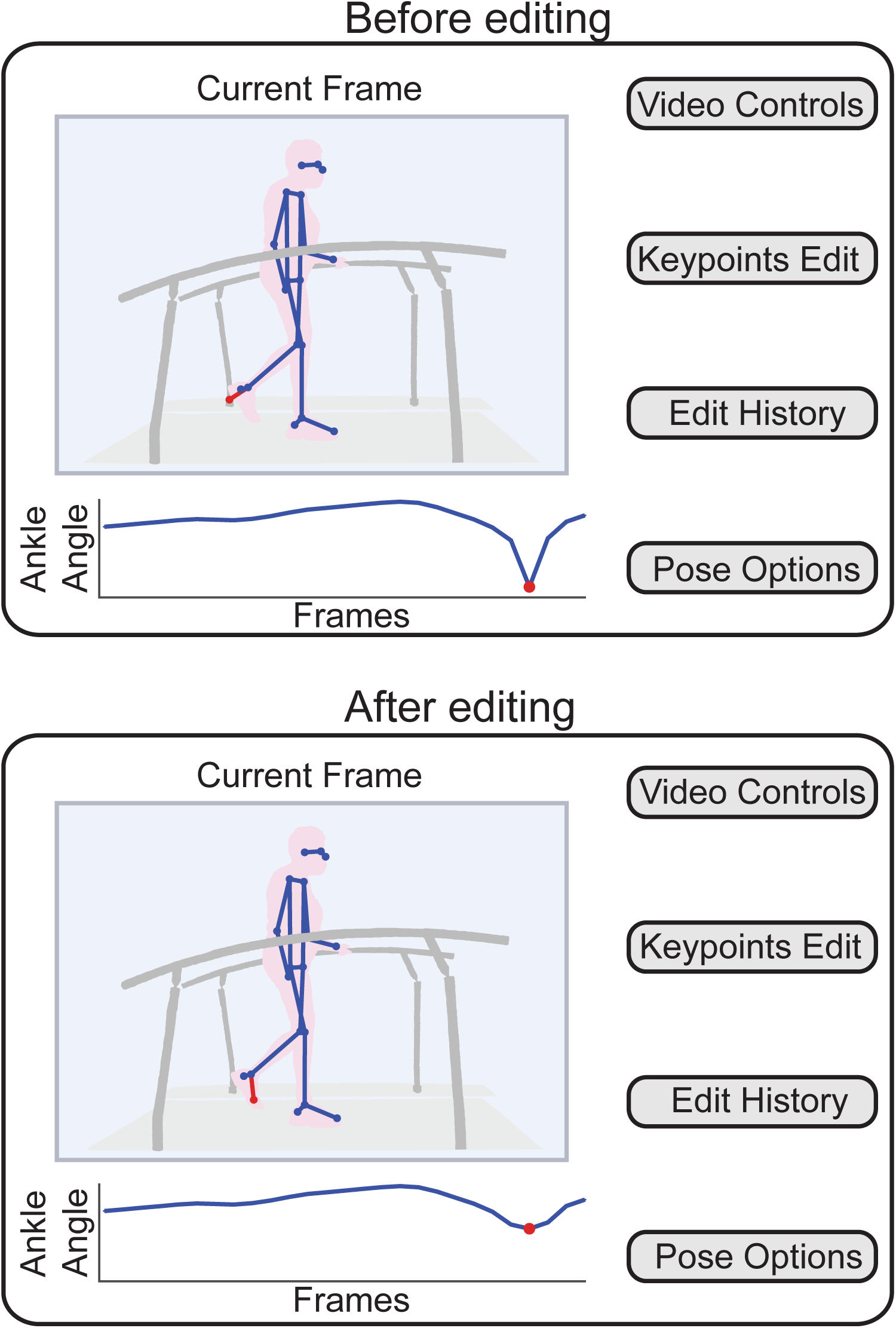
A conceptual schematic of our graphical user interface for manual keypoint editing before (top) and after (bottom) keypoint editing.

These keypoint data can be loaded into the pose editor from a .csv file using the ‘Load Pose (from csv)’ button under the ‘Pose Options’ section of the GUI. This button will prompt the user to select the .csv file that contains the existing keypoint outputs. There is a separate GUI included in the repository to provide additional support for the pose format conversion, (i.e., from OpenPose JSON format to .csv).

#### Human pose estimation within the pose editor GUI

The pose editor also supports the execution of Google Mediapipe (BlazePose [9,25]) locally within the application for subsequent keypoint editing. This can be done on a single frame or across an entire video. The user must first click the ‘Load Video’ button under the ‘Video Controls’ section of the GUI and then select the video that will be used for pose estimation. This will load the selected video into the application, where it can be viewed using the video navigation controls along the bottom edge of the pose editor. For single frame pose estimation, the user then clicks the ‘Run Pose Current Frame’ button under the ‘Pose Options’ section. For full video pose estimation, the user clicks the ‘Run Pose Entire Video’ button within the same section.

#### Video and keypoint display

The pose editor displays the first frame of the video and overlays the corresponding keypoints (frames can be selected using a slider at the bottom of the pose editor). This keypoint overlay is interactive, as the user can either click on a keypoint of interest or use the ‘Select Keypoint’ pulldown menu under the ‘Keypoint Operations’ section to select the desired keypoint. Once selected, the keypoint changes color from green to blue. To facilitate visual inspection of the quality of the keypoints throughout the entire video, time-series plots of x- and y-coordinate trajectories of the selected keypoint are displayed below the video. For the purposes of our evaluation of the pose editor presented in this manuscript, the pose editor also plots the time-series of left and right ankle angles for visual inspection.

The user can also change the appearance of the video frame using the ‘Video Controls’ section allowing the user to load (initial step), rotate, zoom, and toggle the video between color vs. black and white (as black and white can occasionally provide stronger contrast for keypoint editing).

#### Manual keypoint editing

The core functionality of the pose editor is editing mis-tracked or non-tracked (in which the pose estimation model failed to detect the keypoint) keypoints to the correct anatomical locations. The user selects the desired keypoint and manually drags it to the correct location using the cursor. As an example, consider the aberrant tracking of the left foot (big toe) marker in Figure 2 (top). After selecting the keypoint and dragging it to the correct (anatomically realistic under two-dimensional view) location, the edited keypoint location is shown in Figure 2 (bottom). Alternatively, the user can input the values of x- and y-coordinates of a selected keypoint using the ‘X Coordinate’ and ‘Y Coordinate’ controls under the ‘Keypoint Operations’ section and click ‘Update Coordinates.’

Once all desired edits are made to the keypoint data, the user can click the ‘Save Poses (to csv)’ button under the ‘Pose Options’ section to save the edited keypoint coordinates.

### Example application of the pose editor on treadmill walking videos of persons with stroke

#### Participants

We provide a use case of the pose editor GUI using a subset of 23 participants from our previously published data on treadmill walking in chronic stroke because these data were affected by occlusions and aberrant tracking (participants of current study: 15 men, 8 women; age 62±8 years (mean±SD); body mass 87.0±20.1 kg; body height 1.73±0.10 m). Eleven participants used one or more assistive devices during overground walking (10 used a cane, 3 used ankle-foot orthosis, and 1 used functional electrical stimulation of the plantarflexors; no participants used a cane during treadmill walking but participants who used orthoses or functional electrical stimulation also used these devices during treadmill walking). All participants provided written informed consent prior to participation in accordance with the protocol approved by the Johns Hopkins University School of Medicine Institutional Review Board.

#### Protocol and data collection

Participants walked on the treadmill for two minutes at their preferred speeds as determined by overground ten-meter walk tests (they could hold onto the handrails as needed). Occasionally, we slowed the treadmill per participant request. We simultaneously recorded three-dimensional, marker-based motion capture data (detail on the motion capture marker placement [11] and filtering procedures [13] can be found in our previous work) and sagittal video with the camera facing the participant’s right side. For manual editing of the pose estimation keypoints in the current study, we used OpenPose [13].

#### Manual keypoint editing procedure

In our previous study [13], we identified aberrant foot and ankle tracking during several treadmill trials. Here, we used our pose editor to examine the lower extremity keypoints frame-by-frame and manually edit any keypoints which clearly deviated from its anatomical location as determined by visual inspection. We chose to edit keypoints for the initial 300-600 frames of the video. We chose this interval because this yielded a median of eight cycles per analysis, which we considered to be a reasonable trade-off between editing enough gait cycles and a suitable time-commitment for manual inspection. We recorded the time spent editing the keypoints for each participant.

#### Data analysis

After processing the motion capture marker trajectories, we detected left and right heel-strikes and toe-offs as the positive and negative peaks, respectively, of the anterior-posterior left or right lateral malleolus markers relative to the torso to define gait cycles. Gait events were calculated from the motion capture data and OpenPose data independently. We then calculated the sagittal ankle angle separately from 1) motion capture, 2) raw (i.e., unedited) sagittal pose data, and 3) edited pose data for the entire time series. We also interpolated individual gait cycles in time and averaged the sagittal ankle angles across all cycles for each participant to result in a single, normalized ensembled time series within each limb for each participant. This resulted in three datasets of sagittal ankle angles (in both time series and cycle based): ground-truth marker-based motion capture data (MoCap), raw (i.e., unedited) OpenPose data (OP Unedited), and edited OpenPose data (OP Edited). We also recorded the time spent editing the keypoints for each video.

#### Statistical analysis

We conducted several analyses to determine whether the pose editor improved agreement between OpenPose and ground-truth ankle kinematics as measured using a marker-based, three-dimensional motion capture system. First, we calculated 1) mean absolute error (MAE) values to quantify absolute differences between MoCap and the OP Edited and OP Unedited sagittal ankle angles, and 2) Pearson correlation coefficients (r) at zero time-lag between MoCap and OP Edited sagittal ankle angles and between MoCap and OP Unedited sagittal ankle angles to assess relationships between the ground-truth and video-based ankle kinematics before and after keypoint editing. We calculated Pearson correlation coefficients and MAE for each gait cycle per individual (∼3-10 cycles per participant, median=8 cycles, n=23 participants). Individual gait cycle level data were all interpolated to 100 data points per gait cycle to account for lower sampling rates in the OpenPose video data. To compare Pearson correlation coefficients and MAE between OP Edited and OP Unedited methods, we fit separate linear mixed-effects models to account for repeated measures nested within participants and the unbalanced design from different participants contributing varying numbers of gait cycles. Models were fitted separately for right and left sides and included ‘method’ (OP Edited vs. OP Unedited) as a fixed effect, with random intercepts for ‘participant’ and ‘cycle’ to account for between-subject and within-subject cycle-to-cycle variability, respectively. The ‘cycle’ was treated as a random effect because we did not expect systematic trends across gait cycles. Estimated marginal means were calculated for each method, and a pairwise contrast was used to test significant differences between OP Edited and OP Unedited methods.

Next, we performed a modified Bland-Altman analysis with MoCap serving as the ground truth reference to assess the difference, bias, and limits of agreement in peak ankle angles in OP Unedited and OP Edited. This analysis provided a measure of agreement at discrete time points between MoCap and the OP Edited and OP Unedited methods at gait cycle level.

Finally, to assess whether time spent editing keypoints was associated with improvements in tracking accuracy, we used Pearson correlations to examine the relationship between editing time and changes in agreement with ground truth (i.e., differences in MAE and correlation coefficients between OP Unedited and OP Edited with respect to MoCap, averaged first across gait cycles and then across left and right sides). Statistical significance was set at α=0.05. All analyses were conducted using the *lme4* and *emmeans* packages in R.

## RESULTS

### Keypoint editing reduced the error of pose estimation-based sagittal ankle angles relative to ground-truth motion capture

Group-level (mean ± SEM) sagittal right and left ankle angles as estimated by the MoCap, OP Unedited, and OP Edited methods are shown in Figure 3A. We observed significant reductions of mean absolute error (MAE; Figure 3B) for the left and right ankle joint angles after keypoint editing. Right ankle MAE decreased from 5.19°±0.54° (OP Unedited) to 3.85°±0.54° (OP Edited), a mean reduction of 1.34° (95% CI: 0.73-1.95°, t(311)=4.42, p<0.001). Left ankle MAE also showed significant improvement, decreasing from 6.89°±0.42° (OP Unedited) to 4.76°±0.42° (OP Edited), a mean reduction of 2.13° (95% CI: 1.54-2.72°, t(308)=7.18, p< 0.001).

**Figure 3.**
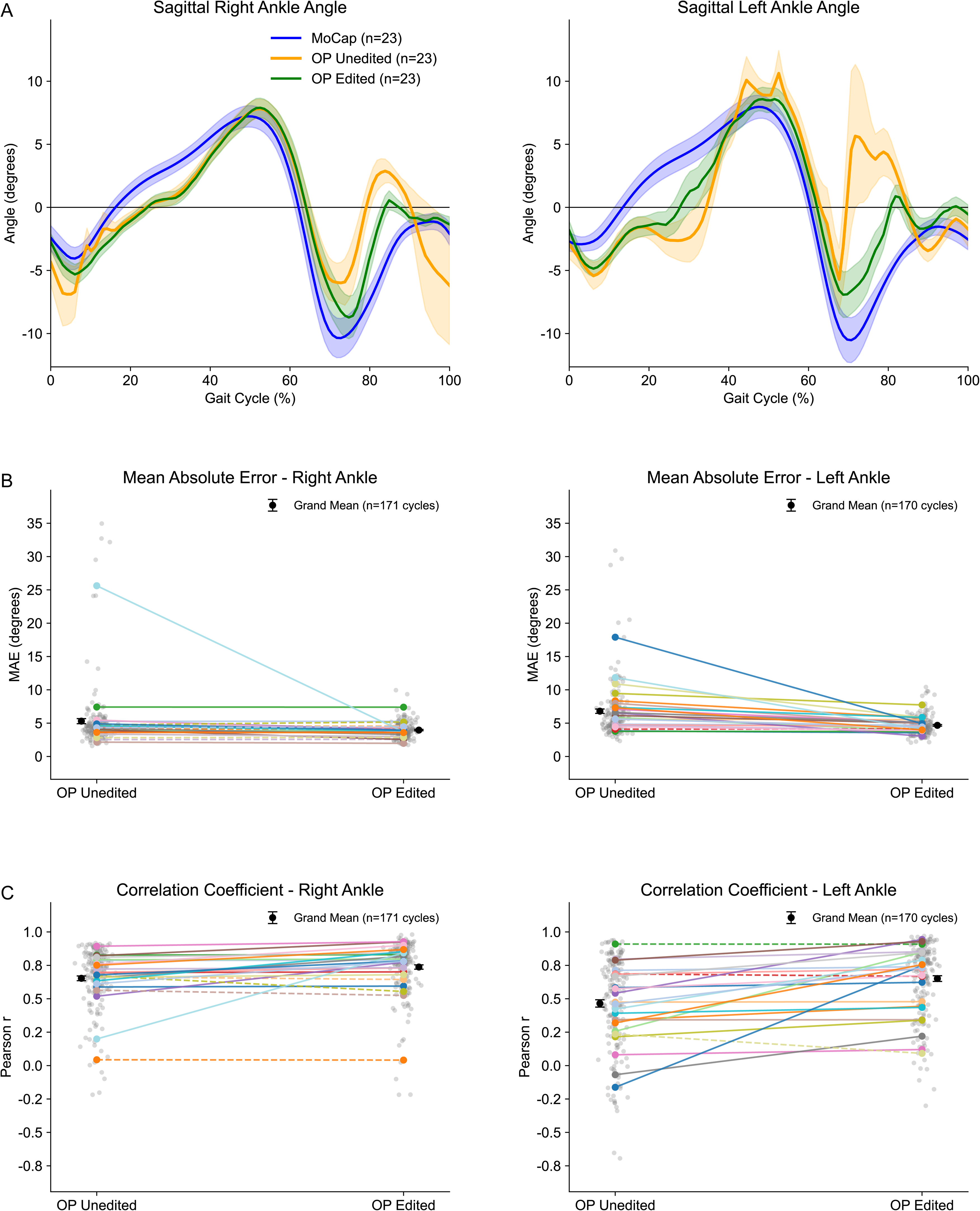
A) Group mean sagittal ankle angles from ground-truth, three-dimensional motion capture data (MoCap), OpenPose keypoints without manual editing (OP Unedited), and OpenPose keypoints after manual editing (OP Edited). B) Mean absolute error (MAE) between OP Unedited and MoCap sagittal ankle angles and between OP Edited and MoCap sagittal ankle angles. C) Pearson’s correlation coefficients (r) at time-lag zero between OP Unedited and MoCap sagittal ankle angles and between OP Edited and MoCap sagittal ankle angles.

### Sagittal ankle angles were more strongly associated with ground-truth motion capture following manual keypoint editing

There were significant improvements in correlation coefficients (r) following editing for both ankle joints (Figure 3C). For the right ankle, coefficients increased from 0.65±0.04 (MoCap vs. OP Unedited) to 0.74±0.04 (MoCap vs. OP Edited), representing a mean improvement of 0.08 (95% CI: 0.05-0.12, t (310)=5.08, p<0.001). The left ankle (i.e., the ankle on the side of the body opposite the camera) demonstrated a larger improvement, with coefficients increasing from 0.44±0.05 to 0.62±0.05, a mean difference of 0.19 (95% CI: 0.14-0.24, t (308)=7.29, p<0.001).

### Sagittal ankle angles demonstrated reduced bias and narrower limits of agreement with ground-truth motion capture following manual keypoint editing

Using a Bland-Altman analysis to assess changes in estimated ankle joint angle minima and maxima (Figure 4), mean differences (MoCap minus OP Unedited or OP Edited) reduced significantly for the right ankle from 4.72° to 0.17° (minimum angles) and from -7.59° to -2.59° (maximum angles). Left ankle measurements showed similar improvements, with mean differences reducing from 9.46° to 2.00° (minimum) and from -18.79° to -4.16° (maximum). For the right ankle, standard deviations decreased from 33.89° to 7.13° (minimum angles) and from 27.28° to 3.84° (maximum angles) when comparing OP Unedited to OP Edited. Left ankle measurements demonstrated similar improvements, with standard deviations reducing from 26.14° to 8.80° (minimum) and from 44.17° to 6.93° (maximum).

**Figure 4.**
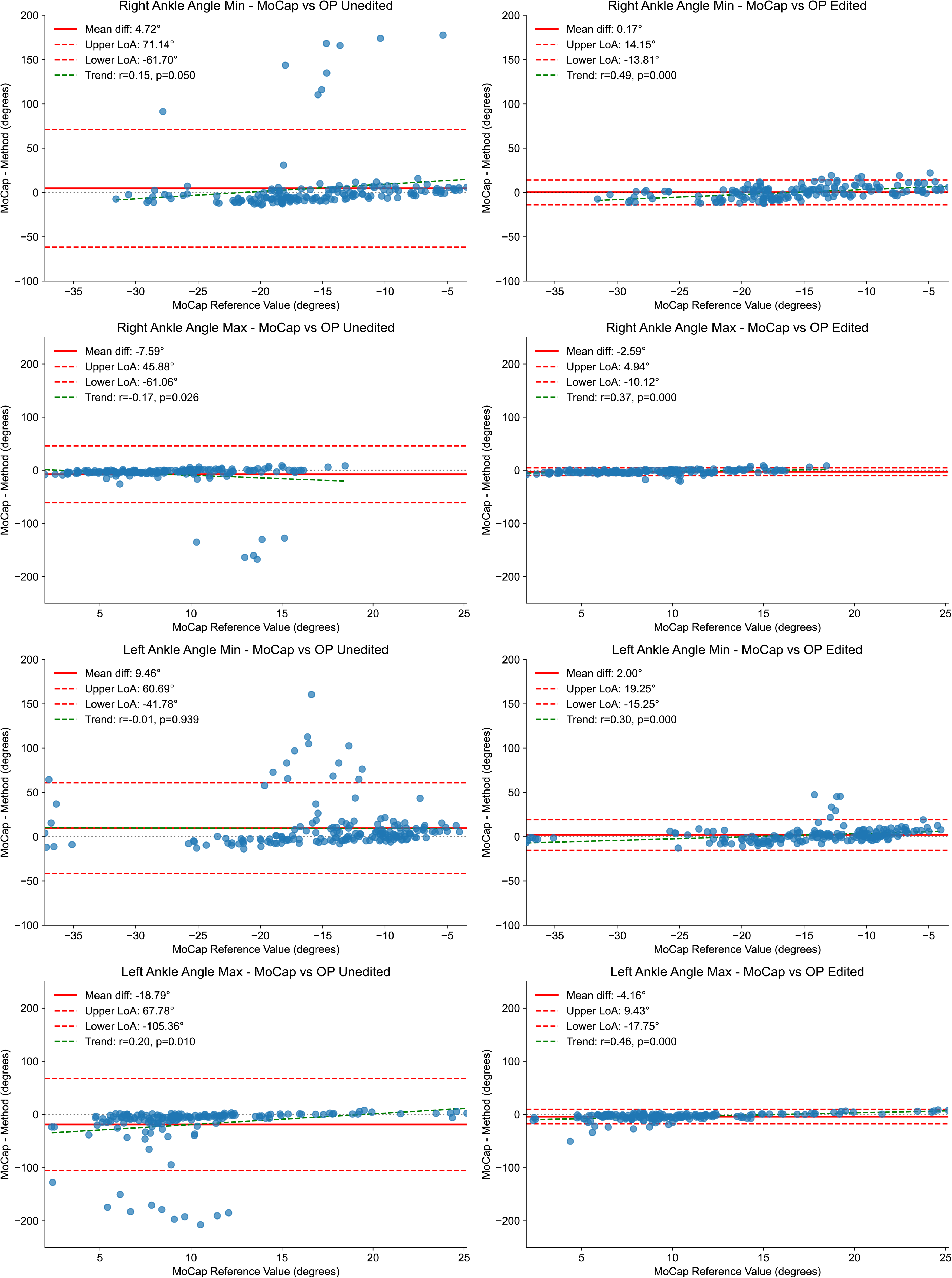
Bland-Altman plots showing distributions of the differences between the video-based and motion capture measurements of right and left sagittal ankle angle minima and maxima against the motion capture-based measurements.

### Time spent on manual keypoint editing was associated with the magnitude of improvement in tracking

We observed a significant negative relationship between time spent editing the keypoints and the mean change in MAE (across both ankles) from OP Edited vs. MoCap vs. OP Unedited vs. MoCap (*r*=-0.69, p<0.001; Figure 5A), indicating that the time spent editing the keypoints was related to larger decreases in MAE. We also observed a significant positive relationship between time spent editing and the mean (i.e., mean of correlation values between the left and right ankles) improvement in Pearson’s correlation coefficient (*r*=0.59, *p*=0.006; Figure 5B), indicating that the time spent editing the keypoints was related to higher correlations between the video-based and MoCap ankle angles.

**Figure 5.**
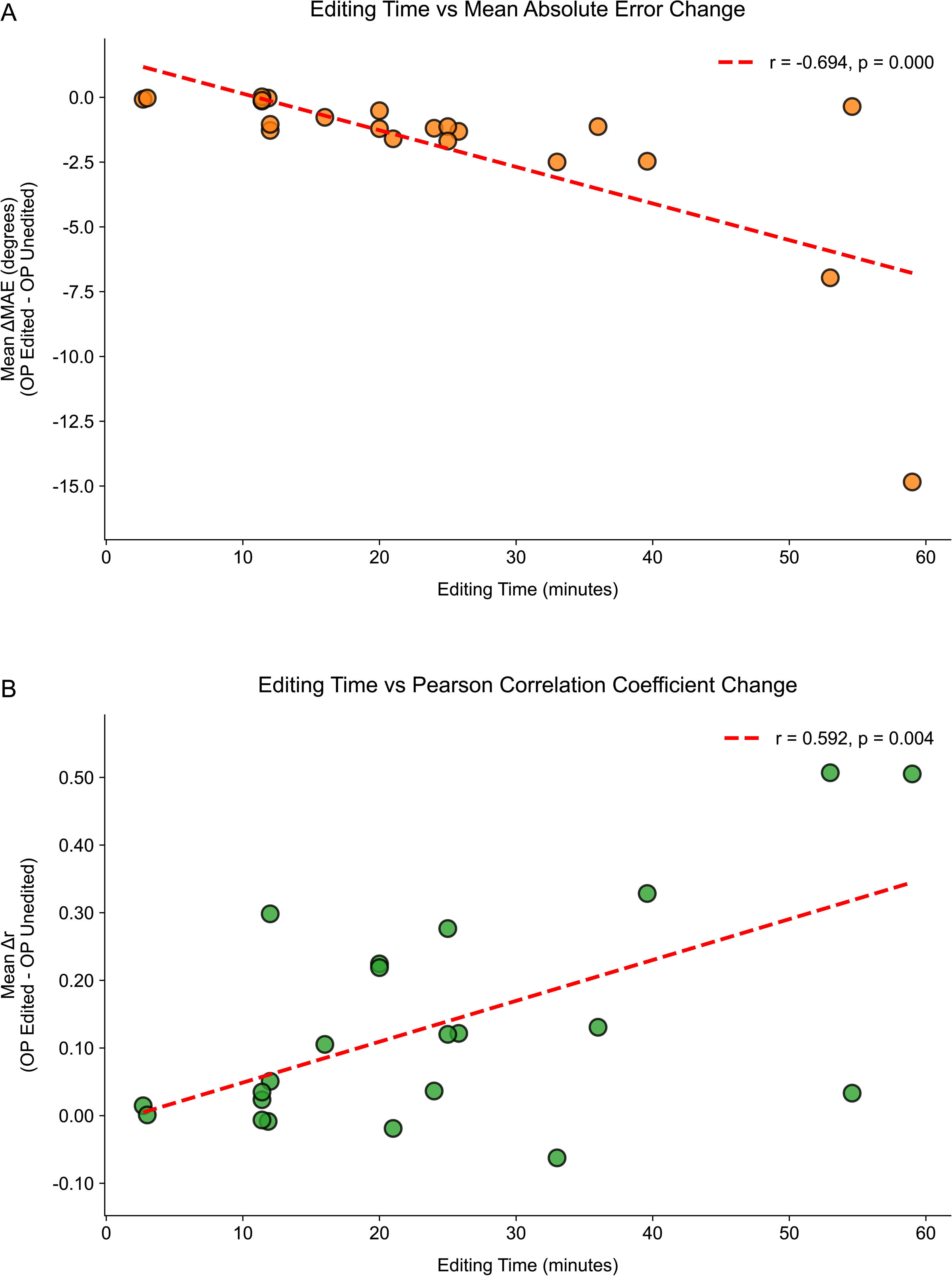
A) Relationships between editing time and mean change in MAE from OP Unedited vs. MoCap to OP Edited vs. MoCap. B) Relationships between editing time and mean change in Pearson correlation coefficients (r) from OP Unedited vs. MoCap to OP Edited vs. MoCap. The red dashed lines represent linear trend lines fit to the data.

## DISCUSSION

Here, we developed and tested a novel GUI for manual keypoint editing for video-based human pose estimation. Specifically, we designed this tool to enable users to fix mis-tracked or untracked keypoints resulting from pretrained human pose estimation algorithms applied to digital videos. The tool provides features for executing a human pose estimation algorithm [9], editing existing keypoints from a previous application of a pose estimation algorithm, editing new keypoints, and observing kinematic results (among others). We also used previously analyzed data from treadmill walking in persons with stroke [13] with known mis-tracked and untracked keypoints to demonstrate a significant improvement in joint angle kinematics following manual editing of the pose estimation keypoints. Finally, we made the pose editor tool freely available on a GitHub repository (https://github.com/JeffZC/pose-editor).

While we applied the pose editor to edit keypoints during videos of treadmill walking, there are many other potential applications. We developed this tool because we and others have observed that pretrained human pose estimation algorithms can result in aberrant movement tracking (as has been summarized in a recent publication [23]), especially in clinical populations and in nonideal settings for video recordings (e.g., settings with poor light, poor contrast between the subject of the video and the environment, etc; [10]). As examples, we have observed that keypoints would occasionally track inanimate objects in the room [13], and collaborators shared that foot keypoints were often inaccurate when participants wore dark shoes when walking on a dark floor [26]. As long as movement can be tracked via video-based pose estimation that results in virtual keypoints, the pose editor could be used to improve the quality of pose estimation-based kinematic measurements in virtually any behavior and in any setting.

Our pose editor tool adds to a growing set of software tools for video-based tracking of human movement kinematics [2–6,8,9,17,27–29]. In addition to algorithms for human and non-human pose estimation [2–6,8,9,28], there are also new tools for flexible implementations of these various algorithms with associated data visualization features [27], mapping of joint trajectories onto interpretable biomechanical outputs [16,17], and translation of video-based kinematics into three-dimensional musculoskeletal modeling [29], among others. The pose editor tool expands capabilities to improve the accuracy of outputs from video-based human pose estimation algorithms by providing the user with more control over the keypoint locations.

In addition to facilitating improved accuracy in kinematics generated by video-based pose estimation algorithms, our pose editor tool also makes it easier for inexperienced users to apply a pretrained model to their videos. We integrated Google Mediapipe directly into the pose editor so that the user has the option to apply pose estimation on either a single frame or whole video with a simple click. We are hopeful that this feature will increase the accessibility of pose estimation to beginning users and also streamline the workflow so that execution of a pose estimation algorithm and subsequent editing of keypoints are possible using a single tool.

As colleagues and collaborators have begun to use the pose editor, we have also garnered useful feedback that will inform future revisions to the tool. We plan to integrate additional pose estimation models into our editor (beyond Mediapipe) so that the users can select their preferred model(s) based on their use case and requirements. We also plan to provide customizability of the kinematic plots so that various joint angles on the body can be plotted to enable the user to identify specific timepoints within the video that may be affected by aberrant tracking. Finally, we will also provide additional software for reformatting existing keypoints output from previous applications of various pose estimation algorithms so that they can be easily brought into the tool for editing.

There are also limitations to using the pose editor. While our tool provides a series of distinctive features for execution of a human pose estimation algorithm and subsequent editing, manual editing can be significantly time consuming. This is particularly limited when the original tracking quality is low and/or when the video is of long duration. We assessed this limitation quantitatively as part of this study, as we found that the degree of improvement in the pose estimation-based kinematics following manual editing was significantly related to the time spent performing the editing. Additionally, although the current iteration of pose editor is limited to executing Google Mediapipe, keypoints resulting from other models can be loaded into the tool if previously generated externally.

## CONCLUSION

We developed and tested a novel tool for execution of a human pose estimation algorithm and manual keypoint editing. Using a previously analyzed dataset of treadmill walking in persons with stroke, we showed that the tool can significantly improve the quality of movement kinematics that result from video-based human pose estimation. We have also made the tool freely available at https://github.com/JeffZC/pose-editor. We hope that this pose editor tool will improve accessibility to clinical and research applications of human pose estimation algorithms and facilitate improved accuracy of resulting movement kinematics.

## Data Availability

All data produced in the present study are available upon reasonable request to the authors.

## ACKNOWLEDGMENTS

We acknowledge funding from the Eunice Kennedy Shriver National Institute of Child Health and Human Development (NIH grant P50HD118624 to RTR, NIH grant T32HD007414 supported RV) and the American Heart Association (grant 23IPA1054140 to RTR). We also thank Dr. Rujuta Wilson, Jeff Anderson, and Anahid Assadourian for providing guidance, testing, and feedback on this project.

