## Supplementary figures and images for "A graphical user interface for editing keypoints from human pose estimation algorithms"

### Supplementary Figure 1

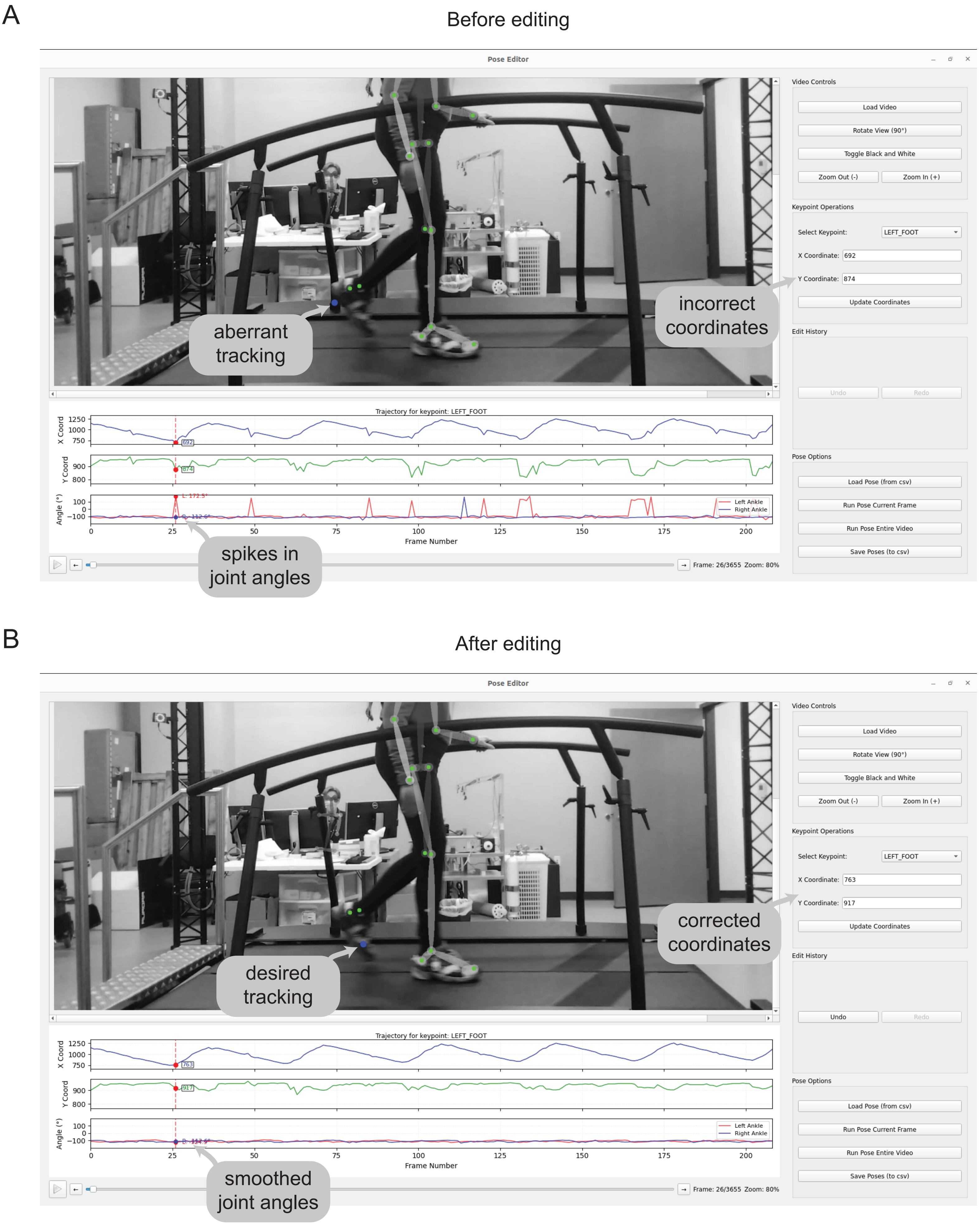
